# Tocilizumab as a Therapeutic Agent for Critically Ill Patients Infected with SARS-CoV-2

**DOI:** 10.1101/2020.06.05.20122622

**Authors:** Russell M. Petrak, Nathan C. Skorodin, Nicholas W. Van Hise, Robert M. Fliegelman, Jonathan Pinsky, Vishal Didwania, Michael Anderson, Melina Diaz, Kairav Shah, Vishnu V. Chundi, David W. Hines, Brian P. Harting, Kamo Sidwha, Brian Yu, Paul Brune, Anjum Owaisi, David Beezhold, Joseph Kent, Dana Vais, Alice Han, Neethi Gowda, Nishi Sahgal, Jan Silverman, Jonathan Stake, Jenie Nepomuceno, Renuka Heddurshetti

## Abstract

**Background:** Tocilizumab is an IL-6 receptor antagonist with the ability to suppress the cytokine storm in critically ill patients infected with SARS-CoV-2.

**Methods:** We evaluated patients treated with tocilizumab for a SARS-CoV-2 infection who were admitted between 3/13/20 and 4/16/20. This was a multi-center study with data collected by chart review both retrospectively and concurrently. Parameters evaluated included age, sex, race, use of mechanical ventilation (MV), usage of steroids and vasopressors, inflammatory markers, and comorbidities. Early dosing was defined as a tocilizumab dose administered prior to or within one (1) day of intubation. Late dosing was defined as a dose administered greater than one (1) day after intubation. In the absence of mechanical ventilation, the timing of the dose was related to the patient’s date of admission only.

**Results:** We evaluated 145 patients. The average age was 58.1 years, 64% were male, 68.3% had comorbidities, and 60% received steroid therapy. Disposition of patients was 48.3% discharged and 29.3% expired, of which 43.9% were African American. Mechanical ventilation was required in 55.9%, of which 34.5% expired. Avoidance of MV (p value = 0.002) and increased survival (p value < 0.001) was statistically associated with early dosing.

**Conclusions:** Tocilizumab therapy was effective at decreasing mortality and should be instituted early in the management of critically ill COVID-19 patients.

**Summary:** Utilizing tocilizumab early in the treatment course of critically ill patients with COVID-19 resulted in significant decreases in mortality and the avoidance of mechanical ventilation.

## Introduction

SARS-CoV-2 is a novel coronavirus, first identified in Wuhan, China in late 2019^1^, that has rapidly expanded to become a worldwide pandemic. Some patients progress to severe illness resulting in hospitalization and respiratory decompensation. Mortality rates have varied from 24.5%-97%^2,3^. The spectrum of symptomatic COVID –19 disease ranges from mild, nonspecific influenza-like symptoms, to a second stage characterized by progressive hypoxia and pulmonary infiltrates. The third and most severe stage of the illness is characterized by rapidly worsening oxygenation, acute respiratory distress syndrome, hemodynamic instability, hypercoagulability, neurologic dysfunction, and myocarditis^4–6^. Multiple cytokines and inflammatory markers such as interleukin (IL)-2, IL –6, C-reactive protein (CRP), ferritin, D-dimer, and lactate dehydrogenase (LDH) frequently become elevated during the final stage of the disease.

While no specific therapy has been identified and validated in clinical trials, tocilizumab has been suggested as a possible immunomodulatory agent to offset what has been referred to as the cytokine storm^7^. Arresting this process is thought to be key in reducing morbidity and mortality in this patient population^8,9^. As an IL-6 receptor antagonist, tocilizumab was originally approved in 2010 and has indications for the treatment of rheumatoid arthritis and the cytokine release syndrome (CRS). Its approval for use in CRS provides rationale for the treatment of severe COVID-19 disease^10–11^.

Current tocilizumab dosing for COVID-19 has been 4–8 mg/kg (max: 800mg per dose) intravenous infusion, repeated once if necessary^11–13^.

Due to the ongoing urgency of the COVID-19 pandemic, this study was designed to identify the utility and timing of tocilizumab dosing for critically ill patients infected with SARS-CoV-2.

## Methods

This study was conducted by Metro Infectious Disease Consultants (MIDC), a fully integrated infectious disease (ID) private practice composed of 108 ID physicians. This was a multi-center study evaluating patients treated with tocilizumab for a SARS-CoV-2 infection who were admitted between 3/13/20 and 4/16/20. Patients were evaluated retrospectively and concurrently through chart review and direct interactions with the prescribing MIDC physician. This study was independently approved by Western IRB. Informed consent was deemed unnecessary because all personal data was de-identified prior to the analysis. Tocilizumab was prescribed at the discretion of the ID physician with consideration of local hospital treatment protocols. All patients treated with tocilizumab were included regardless of age, race, or risk factors. Patients were excluded if data was unavailable for review. Parameters for evaluation included the patient’s age, sex, race, date of admission, length of hospital stay (LOS), use of mechanical ventilation, usage of steroids, usage of vasopressors, hydroxychloroquine (HCQ) and azithromycin (AZ) in combination, and remdesivir therapy. Comorbidities evaluated included age greater than sixty (60) years old, diabetes, chronic obstructive pulmonary disease, bronchospastic illness, chronic cardiac or renal disease, immunodeficiency or neoplastic disease. The tocilizumab dose was recorded by number of doses administered, dosing regimen, and the timing of administration in relation to the date of hospital admission and mechanical ventilation (MV). Early dosing was defined as a tocilizumab dose administered prior to or within one (1) day of intubation. Late dosing was defined as a dose administered greater than one (1) day after intubation. In the absence of mechanical ventilation, the timing of the dose was related to the patient’s date of admission only. Markers of inflammation including ferritin, D-dimer, LDH, and CRP were recorded at the time of dosing. Patient dispositions were recorded as discharged from the hospital, expired, or continued hospitalization.

## Statistical Analysis

Descriptive statistics for baseline patient characteristics, clinical variables, and outcomes were summarized using means and standard deviations for continuous variables and counts and percentages for categorical variables. Continuous outcomes were evaluated using t-tests. Chi-squared tests or Fisher Exact tests were used to evaluate categorical outcomes. Potential confounding variables including age, gender, race, and comorbidities were adjusted for using propensity score methods. Propensity scores, defined as the probability of being in either intervention group, were calculated from a regression model with the dose timing (early vs late) as the outcome and baseline demographic and clinical variables of each participant as predictor variables. To adjust the sample for these baseline confounding variables, the propensity scores were converted to weights (“inverse probability of treatment weights”, or IPTW), and the outcomes were then compared using weighted versions of the Chi-Square test or Fisher’s exact test for categorical outcomes or a weighted version of the t-test for continuous outcomes. P-values for both the unadjusted and weighted versions of the tests are reported.

To confirm robustness of the propensity score weighted test results, multivariable regression models were also fit for each outcome, adjusting for baseline patient demographic variables. Statistical significance for all methods was defined as a p-value ≤ 0.05.

## Results

One hundred fifty-seven hospitalized patients were evaluated from 24 hospitals in 4 states and were treated by 26 individual MIDC physicians (Table 1). Twelve patients were excluded from analysis secondary to incomplete records. The average age was 58.1 years and did not vary significantly between those who were discharged, expired, required mechanical ventilation, or received steroids. Ninety-three (64.6%) of the patients were male. Fifty-six (41%) patients were White, 43 patients (29.7%) were African American, 32 were Hispanic (22.1%), and 5 (3.5%) were Asian. Comorbidities were present in 99 (68.3%) patients. Eighty-seven (60%) patients received steroid therapy, and 143 (98.6%) received HCQ and AZ in combination at standard doses. Inflammatory markers were elevated in all patients. Mechanical ventilation was required in 81 (55.9%) of patients. The LOS for all discharged patients was 15.3 days, expired patients was 16.3 days, and those remaining in the hospital at the time of data analyses is 25.8 days. Table 1 shows the average baseline patient demographics and clinical characteristics. Both the unadjusted and propensity score weighted results show no significant differences in any of the demographic data. D-dimer was the only clinical parameter found to be significant between patients who received the tocilizumab dose early vs. late in their treatment course.

**Table 1.**
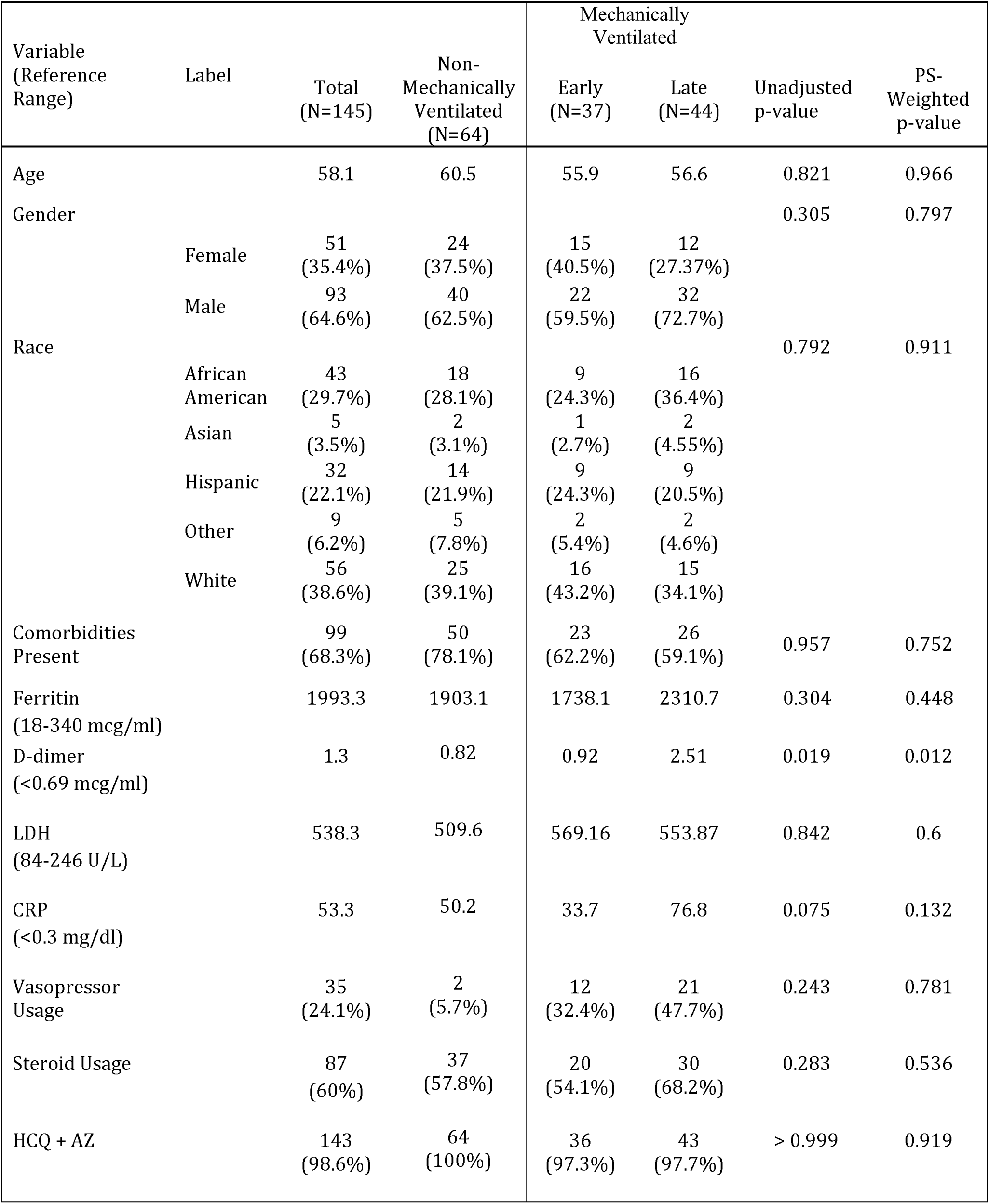
Demographics and Clinical Characteristics.

| Variable<br>(Reference<br>Range) | Label | Total<br>(N=145) | Non-Mechanically<br>Ventilated<br>(N=64) | Mechanically<br>Ventilated |  | Unadjusted<br>p-value | PS-Weighted<br>p-value |
| --- | --- | --- | --- | --- | --- | --- | --- |
|  |  |  |  | Early<br>(N=37) | Late<br>(N=44) |  |  |
| Age |  | 58.1 | 60.5 | 55.9 | 56.6 | 0.821 | 0.966 |
| Gender |  |  |  |  |  | 0.305 | 0.797 |
|  | Female | 51<br>(35.4%) | 24<br>(37.5%) | 15<br>(40.5%) | 12<br>(27.37%) |  |  |
|  | Male | 93<br>(64.6%) | 40<br>(62.5%) | 22<br>(59.5%) | 32<br>(72.7%) |  |  |
| Race |  |  |  |  |  | 0.792 | 0.911 |
|  | African<br>American | 43<br>(29.7%) | 18<br>(28.1%) | 9<br>(24.3%) | 16<br>(36.4%) |  |  |
|  | Asian | 5<br>(3.5%) | 2<br>(3.1%) | 1<br>(2.7%) | 2<br>(4.55%) |  |  |
|  | Hispanic | 32<br>(22.1%) | 14<br>(21.9%) | 9<br>(24.3%) | 9<br>(20.5%) |  |  |
|  | Other | 9<br>(6.2%) | 5<br>(7.8%) | 2<br>(5.4%) | 2<br>(4.6%) |  |  |
|  | White | 56<br>(38.6%) | 25<br>(39.1%) | 16<br>(43.2%) | 15<br>(34.1%) |  |  |
| Comorbidities<br>Present |  | 99<br>(68.3%) | 50<br>(78.1%) | 23<br>(62.2%) | 26<br>(59.1%) | 0.957 | 0.752 |
| Ferritin<br>(18-340 mcg/ml) |  | 1993.3 | 1903.1 | 1738.1 | 2310.7 | 0.304 | 0.448 |
| D-dimer<br>(<0.69 mcg/ml) |  | 1.3 | 0.82 | 0.92 | 2.51 | 0.019 | 0.012 |
| LDH<br>(84-246 U/L) |  | 538.3 | 509.6 | 569.16 | 553.87 | 0.842 | 0.6 |
| CRP<br>(<0.3 mg/dl) |  | 53.3 | 50.2 | 33.7 | 76.8 | 0.075 | 0.132 |
| Vasopressor<br>Usage |  | 35<br>(24.1%) | 2<br>(5.7%) | 12<br>(32.4%) | 21<br>(47.7%) | 0.243 | 0.781 |
| Steroid Usage |  | 87<br>(60%) | 37<br>(57.8%) | 20<br>(54.1%) | 30<br>(68.2%) | 0.283 | 0.536 |
| HCQ + AZ |  | 143<br>(98.6%) | 64<br>(100%) | 36<br>(97.3%) | 43<br>(97.7%) | > 0.999 | 0.919 |

## Treatment

One hundred twenty-three (84.8%) patients received a single dose of tocilizumab and 22 (15.2%) received two doses separated by twelve hours. One hundred thirty-five (135) patients received a dose of four (4) milligrams per kilogram (mg/kg) to a maximum dose of 400 mg, 5 patients received a single dose of 600 mg and 4 patients a dose of 800 mg. Of the MV patients, 37 received tocilizumab early and 44 late.

## Outcomes

Seventy patients (48.3%) were discharged and 34 patients (23.5%) remain hospitalized. Forty-one patients (28.3%) expired, of which 18 were African American (43.9%), 11 White (26.8%), and 10 Hispanic (24.4%). Eighteen expired patients were over the age of sixty (43.9%). Comorbidities were present in 25 (60.9%) expired patients and 35 (85.3%) required MV. Six patients who expired were not intubated secondary to living will directives.

Table 2 shows the results of a logistic regression model with ventilator usage as the outcome and days from admission to tocilizumab dose, steroid usage and demographics as predictor variables. For each additional day that the tocilizumab dose is delayed from the day of admission, the odds of requiring MV increase by 21%, holding all other covariates constant (95% CI: [1.08, 1.38], p = 0.002). No significant difference was demonstrated on the likelihood of MV related to the use of steroids (p = 0.965).

**Table 2.**
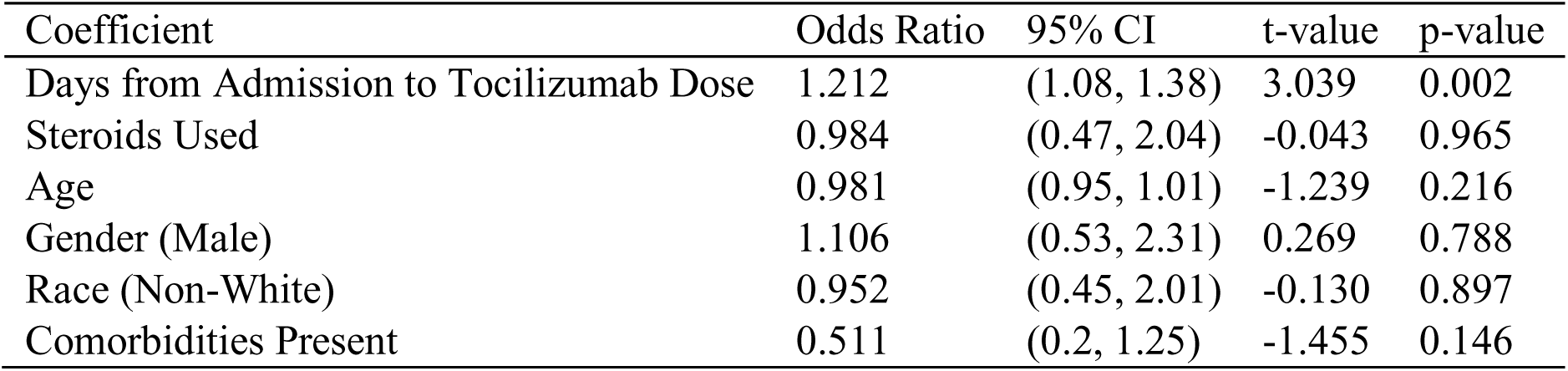
Logistic Regression with Ventilator Status as the Outcome.

| Coefficient | Odds Ratio | 95% CI | t-value | p-value |
| --- | --- | --- | --- | --- |
| Days from Admission to Tocilizumab Dose | 1.212 | (1.08, 1.38) | 3.039 | 0.002 |
| Steroids Used | 0.984 | (0.47, 2.04) | -0.043 | 0.965 |
| Age | 0.981 | (0.95, 1.01) | -1.239 | 0.216 |
| Gender (Male) | 1.106 | (0.53, 2.31) | 0.269 | 0.788 |
| Race (Non-White) | 0.952 | (0.45, 2.01) | -0.130 | 0.897 |
| Comorbidities Present | 0.511 | (0.2, 1.25) | -1.455 | 0.146 |

Table 3 shows the primary outcomes by the timing of tocilizumab dosing in patients requiring MV. A significantly lower mortality rate is demonstrated in the patients who received the tocilizumab dose earlier as compared to those who received the dose later (13.5% vs. 68.2%, p < 0.001), after adjusting for baseline demographic variables. Patients who received the dose earlier had a significantly higher discharge rate compared to those who received the dose later (59.5% vs. 18.2%, p < 0.001).

**Table 3.** Outcomes by Timing of Tocilizumab Dosing.

| Variable | Early<br>(N=37) | Late<br>(N=44) | Unadjusted p-value | PS-Weighted p-value |
| --- | --- | --- | --- | --- |
| Mortality | 5 (13.5%) | 30 (68.2%) | < 0.001 | < 0.001 |
| Discharged | 22 (59.5%) | 8 (18.2%) | < 0.001 | < 0.001 |
| Length of Stay<br>(Days) | 15.85 | 18.87 | 0.162 | 0.354 |

The 30 discharged patients who required MV received the dose early, within one day of intubation, and an average of 4.2 days after admission. Patients who expired received the dose late, an average of 4.1 days after intubation, and an average of 5.4 days after admission.

Table 4 shows the relationship between the timing of tocilizumab dosing and the likelihood that intubated patients would be discharged or expire. The only significant variable predicting that a patient requiring MV would be discharged was the time from intubation to dosing (p < 0.001).

**Table 4:** Effect of Dose Timing between Discharged MV Patients and Expired MV Patients.

| Variable | Label | MV Discharged<br>(N=30) | MV Expired<br>(N=35) | p-value |
| --- | --- | --- | --- | --- |
| Age |  | 56 | 54.5 | 0.639 |
| Gender |  |  |  | 0.221 |
|  | Female | 12 (40%) | 8 (22.9%) |  |
|  | Male | 18 (60%) | 27 (77.1%) |  |
| Race |  |  |  | 0.052 |
|  | African American | 6 (20%) | 16 (45.7%) |  |
|  | Asian | 2 (6.7%) | 1 (2.9%) |  |
|  | Hispanic | 6 (20%) | 10 (28.6%) |  |
|  | Other | 2 (6.7%) | 1 (2.9%) |  |
|  | White | 14 (46.7%) | 7 (20%) |  |
| Comorbidities Present |  | 17 (56.7%) | 19 (54.3%) | > 0.999 |
| Days from Admission to<br>Tocilizumab Dose |  | 4.2 | 5.5 | 0.091 |
| Steroid Usage |  | 15 (50%) | 25 (71.4%) | 0.13 |
| Days from Intubation to<br>Tocilizumab Dose |  | 1.1 | 4.1 | < 0.001 |

Table 5 shows the results of a logistic regression model of patients on mechanical ventilation with mortality as the outcome and dosage timing as well as demographics as predictor variables. The odds of mortality are 17.8 times higher for patients receiving the dose of tocilizumab later compared to patients receiving the dose earlier, holding all other covariates constant (95% CI: [5.32, 74.55], p < 0.001). The odds of mortality are 6.01 times higher for nonwhite patients receiving the dose earlier as compared to white patients receiving the dose earlier, holding all other covariates constant (95% CI: [1.65, 25.66], p = 0.009). For patients who were not placed on mechanical ventilation, the time from admission to dosing was 3.4 days for white and 3.5 days for non-white patients. Six non-intubated patients expired secondary to living will directives.

**Table 5.** Logistic Regression with Mortality as the Outcome.

| Coefficient | Odds Ratio | 95% CI | t-value | p-value |
| --- | --- | --- | --- | --- |
| Late Timing | 17.758 | (5.32, 74.55) | 4.332 | < 0.001 |
| Age | 1.001 | (0.94, 1.06) | 0.037 | 0.971 |
| Gender (Male) | 2.416 | (0.69, 9.23) | 1.349 | 0.177 |
| Race (Non-White) | 6.009 | (1.65, 25.66) | 2.594 | 0.009 |
| Comorbidities Present | 0.597 | (0.12, 2.74) | -0.660 | 0.509 |

Of the patients treated with steroids, 38 (43.7%) were discharged, 30 (34.5%) expired, and 19 (21.8%) remain in the hospital. Fifteen discharged patients received steroids and required MV, compared to 25 patients who expired. The average time from admission to tocilizumab dosing for patients who also received steroids was 4.3 days for discharged patients and 5.7 days for patients who expired.

Of the patients who received two doses of tocilizumab, 11 (50%) were mechanically ventilated, 10 (45.5%) were discharged, 5 (22.7%) expired, and 7 (31.8%) remain in the hospital.

## Discussion

This multi-center, multi-state, observational study displays a real-time evaluation of tocilizumab usage in community hospitals.

The data presented represents, to the best of our knowledge, the largest group of SARS-CoV-2 infected patients treated with tocilizumab in a private practice setting. Our patients were slightly younger than several previous reports at 58.1 years old^2,3^’ Males represented 64% of the patient population and African American patients experienced the highest mortality rate (43.9%), similar to previous reports^14^.

Overall, 28.3% of the patients expired. A statistically significant mortality differential was characterized by the timing of the tocilizumab administration. Patients on mechanical ventilation who received tocilizumab within several days of admission or the day of intubation were more likely to be discharged (p = < 0.001). Patients who received the dose later than one day after intubation were approximately eighteen times more likely to expire (p = < 0.001). As has been delineated in other studies^14^, non-White patients on mechanical ventilation were 6 times more likely to expire despite insignificant differences in age, gender, and comorbidities (p = 0.009). Excluding patients who were not intubated secondary to withdrawal of medical care, 7/52 (13.5%) white patients and 28/87 (32.2%) non-white patients expired. Further studies need to define this repeatedly identified and poorly defined predisposition toward severe and ultimately fatal illness in the non-white population.

Critically ill patients with COVID –19 who require MV have predictably increased rates of mortality that have varied widely in previous reports^2,3,15^. Mechanical ventilation was required in 56% of our patients, with a mortality rate of 42.7%, less than some previous publications. Our study shows that the likelihood of requiring MV increased by 21% for every day the tocilizumab therapy was delayed. Non-intubated patients received the dose approximately one day earlier than the age, sex, comorbidity matched, and ventilated counterparts. This relationship suggests that earlier tocilizumab dosing may prevent the need for MV which will need to be defined by further studies.

The etiology of the delay in tocilizumab therapy for individual patients was difficult to define. Verbally acknowledged reasons included the lack of drug availability, protocol requirements that had not been fulfilled, or lack of consensus between consulting physicians on the efficacy of therapy. Regardless, a delay in dosing resulted in an increased mortality and need for mechanical ventilation, regardless of age, race, or the presence of comorbidities.

The patients who received two doses had similar dispositions compared to those who received a single dose. The small sample size makes any conclusions difficult. Further studies should define the optimal tocilizumab dose and interval.

Steroids were not found to affect patient disposition or the likelihood of MV. Only 60% of patients overall and 32% of those eventually discharged received this therapy. Recommendations for the use of steroids evolved over the study period which may have affected the results. Further studies will be needed to define the type, dose, duration, and timing of this therapy as an adjunct to tocilizumab, or as a stand-alone intervention. The vast majority of patients received HCQ and AZ therapy, suggesting no clinical influence on outcome or need for mechanical ventilation. While the study was not designed to identify the toxicity of this combination, no adverse reactions were documented.

The relationship between inflammation and a prothrombotic state induced by SARS-CoV-2 is evolving^16^. As was seen in our patient population, D-dimer is frequently elevated and may portend a poor prognosis^17^. In addition, IL-6 induces thrombin generation and is correlated with increased fibrinogen levels^18^. While our study was not designed to evaluate the effect of tocilizumab on the procoagulant state, further studies may delineate this as a secondary benefit.

Our study has certain limitations. This study was observational and no control group was utilized for comparison. We did not evaluate the relationship between IL-6 levels and tocilizumab efficacy as these levels were infrequently available at the time of dosing. We did not identify any serious adverse drug reactions, but reactivation of hepatitis B or latent tuberculosis were not formally evaluated. As with all critically ill patients, multiple therapeutic modalities are implemented concurrently in an attempt to minimize morbidity and mortality. Similarly, the patients who received tocilizumab were also treated with antibiotics, anticoagulants, and steroid therapy in varying combinations. Accordingly, it is difficult to identify the most clinically influential modality. Further randomized studies will be needed to answer this question.

In the absence of definitive data, this study should serve as a guidepost to those striving to identify the utility of tocilizumab in the therapeutic armamentarium. While the optimal time to dose tocilizumab has not been previously established, our data strongly supports a mortality benefit of dosing tocilizumab early and within 1 day of intubation. Accordingly, we strongly encourage the use of this agent earlier in the COVID-19 treatment spectrum.

## Data Availability

Upon request, we will make a de-identified dataset available

## Acknowledgements

We thank Farrin A. Manian, MD, MPH, FACP, FIDSA, FSHEA of Massachusetts General Hospital and Harvard Medical School for his timely and critical review of the manuscript. We thank Caleb Scheidel, PhD and Jeremy Albright, PhD of MethodsConsultants for their timely and accurate presentation of our statistical analyses. We would also like to thank all of our esteemed pulmonologists and critical care specialists who tirelessly co-treated these patients with us, without whom this data could not have been collected.

## Notes

### Competing Interest Statement

The authors have declared no competing interest.

### Funding Statement

No external funding was received

### Author Declarations

Western IRB considered this an exempt project

